# Descriptive epidemiology of COVID-19 outcomes in England, from September 2023 to April 2024

**DOI:** 10.1101/2024.11.12.24317146

**Authors:** Nurin Abdul Aziz, Hester Allen, Kiara Assaraf, Gavin Dabrera, Mary Ramsay, Alex Allen

**Affiliations:** Immunisation and Vaccine Preventable Diseases Division, UK Health Security Agency, 61 Colindale Avenue, London, NW9 5EQ, UK

## Abstract

**Introduction:** COVID-19 disease has been associated with severe illness, hospitalisation and death, however, widespread vaccination coverage in England has resulted in reduced disease severity. From 2022, the national vaccination programme has been run twice per year, prioritising older age groups or those classified as clinically vulnerable. Here we assess the trends in COVID-19 outcomes between September 2023 and April 2024, using national-level data held by the UK Health Security Agency (UKHSA).

**Methods:** Data linkage of national-level COVID-19 episode data, NHS emergency and hospital attendance data, and death registrations were used to analyse COVID-19 outcomes. Outcomes were defined as COVID-19 associated A&E attendances, hospital admissions, severe hospitalisations, and deaths.

The number and rate of each COVID-19 outcome category between September 2023 and April 2024 was calculated, stratified by clinical risk status and age and sex.

**Results:** The most common COVID-19 outcomes during this time-period were A&E attendance and hospital admission, with the rates highest among those aged 75 and over. Among this age group, all outcomes disproportionately affect those who have been identified as at clinical risk and those who were immunosuppressed.

High rates of A&E attendance and hospital admission were also observed among infants (under 6 months old) but were lower for more severe outcomes.

**Discussion:** Groups that were most affected by COVID-19 outcomes were currently prioritised for COVID-19 vaccination in England, which will help protect against more severe outcomes including admission to intensive care and death.

Routine national levels surveillance of COVID-19 outcomes is essential to monitor populations most of severe disease and informing vaccination policy.

## Background

COVID-19 has been associated with severe disease, resulting in hospitalisation and death. Older individuals and those who are immunosuppressed or have underlying health conditions have been most impacted by severe COVID-19 disease.

There has been evidence of effectiveness of COVID-19 vaccines against COVID-19 hospitalisations and deaths, following the roll out of the vaccination programme in late 2020 (1–3). The initial rollout included a primary course of COVID-19 vaccine offered to all individuals aged 18 and over in England, followed by a booster dose offered from November 2021 (4). Since then, the national vaccination programme has been run biannually, providing routine boosters prioritising older age groups or those classified as clinically vulnerable.

Through the pandemic, trends in COVID-19 outcomes have changed alongside increased population immunity due to widespread vaccination coverage and previous infection, and the emergence of SARS-CoV-2 lineages that have been associated with less severe disease than previous lineages (5–7).

In this analysis, we assess the temporal and demographic trends in COVID-19 severity between September 2023 and April 2024, which coincides with the Autumn 2023 vaccine programme, using national-level data held by the UK Health Security Agency (UKHSA). Here, we indicate recent trends in those who experience severe COVID-19 disease to understand populations that are most likely to benefit from further boosters of COVID-19 vaccination.

## Methods

### Data sources and linkage

To assess trends in COVID-19 severity, national-level data for COVID-19 episodes, NHS accident and emergency (A&E) attendances and hospital admissions, and registered deaths were utilised.

COVID-19 cases in England are monitored using an episode-based definition to include possible reinfections(8). Each infection episode, beginning with the earliest positive test date, is counted separately if there are at least 91 days between positive test results. Individual-level data for positive COVID-19 episodes were obtained from the Second-Generation Surveillance System (SGSS). SGSS is a national laboratory reporting system in England that captures routine laboratory data on infectious diseases, including positive SARS-CoV-2 specimens, submitted via diagnostic laboratories in compliance with Health Protection (Notification) Regulations (2010)(9,10). These data include information on demographic characteristics of individuals with a positive COVID-19 episode.

As part of routine surveillance, positive COVID-19 episode data from SGSS is routinely enhanced using linkage to the NHS spine through Demographic Batch Service (DBS) tracing to add and validate patient identifiers, including the NHS number unique identifier.

Data on A&E attendances and hospital admissions, including admission to intensive care units (ICU), were obtained from the Emergency Care Data Set (ECDS) and Secondary Users Service (SUS) data set respectively(11,12). These are both national administrative datasets operated for commissioning NHS services that capture information on all activity in NHS A&E departments and NHS hospital and ICU admissions in England.

Individual-level death records, including information on cause of death, were obtained from the national death registration data for England and Wales collated by the Office for National Statistics (ONS)(13,14).

COVID-19 episodes with a specimen date between 21 August 2023 and 30 April 2024 were used for linkage to ECDS and SUS data to provide enough follow-up time following a positive test for a hospital attendance or admission starting on 1 September 2023. Hospital episodes between 1 September 2023 to 30 April 2024 were included in this report.

COVID-19 episodes records were linked to ECDS, SUS and the death registration data on an individual level, linking on the NHS number.

Clinical risk status for individuals with a COVID-19-associated outcome is derived from to NHS Cohorting as a Service (CaaS) data(15), an NHS England service which uses existing individual electronic health records, including primary and secondary health care records, to identify people in clinical risk groups. Risk groups are defined in the COVID-19 Green Book (4) as those with underlying health conditions that may put them at higher risk of severe COVID-19 disease. This includes a range of groups including those who are immunosuppressed, and those with other underlying health conditions without immunosuppressions (referred to as “clinical risk but not immunosuppressed”). Persons with a valid NHS number for linkage but without a record of a clinical risk were classified as “not at clinical risk”.

Clinical risk status was also aggregated by wider under-75 and 75-and-over age bands to account for eligible groups for vaccination. For the most recent Spring 2024 campaign, the eligible groups were all those 75 years and older, and those aged 6 months to 74 years who were immunosuppressed. Data on clinical risk status is as of 1 September 2023.

### Outcome definitions

To assess the severity of COVID-19 during this time-period, we quantify the number of COVID-19 associated outcomes including A&E attendances, hospital admissions, severe hospitalisations, and deaths in England.

Here, COVID-19 A&E attendances were defined as any attendance in A&E up to 14 days after, or 1 day before the earliest positive test of a COVID-19 episode where a respiratory SNOMED diagnosis was recorded.

Similarly, COVID-19 hospital admissions were defined as any hospital admission up to 14 days after or 1 day before the earliest positive test where the admission record was associated with a respiratory ICD-10 diagnosis code. Hospital admissions were also included where the record was associated with a COVID-19 ICD-10 diagnosis code if no record of a positive test was captured in SGSS.

Not all individuals admitted to hospital will have a record of A&E attendance in the study data set. Some hospital admissions occur directly via general practitioners or consultants in ambulatory clinics. Additionally, data on A&E attendances and hospital admissions do not always align on an individual level as they come from two different national data sets. Due to differing coding practices between A&E and inpatient departments, the definition for COVID-19-associated A&E attendance does not include those that had no associated test, whereas the definition for COVID-19 hospital admissions used in this analysis includes individuals with COVID-19 diagnoses indicated by diagnostic codes with no associated tests. This may contribute to differences in numbers between those attending A&E and those admitted to hospital.

COVID-19 hospitalisations were defined as severe if SUS data indicated the length of stay was 2 or more days, and the main specialty code or treatment function related to intensive care medicine or ventilation use or oxygen use.

COVID-19 deaths were defined as deaths where COVID-19 was mentioned as a cause of death on the death certificate. Cause of death is determined through clinical judgment and therefore a positive COVID-19 test record is not required to indicate a person’s death is related to COVID-19.

### Analysis

We present general trends in COVID-19 epidemiology, including crude incidence rates per 100,000 and test positivity, followed by trends in COVID-19 outcomes occurring between 1 September 2023 and 30 April 2024.

For each outcome, we calculate the incidence rate per 100,000 population, stratifying by age, sex, and clinical risk status. Population estimates used for the incidence rates were derived from the mid-year 2021 population estimates from ONS (16).

Results are presented at outcome event-level; hence, individuals may be represented multiple times if they have had more than one event occur for their COVID-19 infection.

## Results

### Epidemiology summary

From 21 August 2023 to 30 April 2024, there were 216,145 total COVID-19 episodes captured in SGSS. Episodes peaked at the end of September and the start of October 2023 at over 2,500 new episodes per day, and reached a nadir from February 2024 to April 2024 where the number of episodes remained relatively low, below 400 per day.

COVID-19 test positivity fluctuated between 17.6% and 4.2%, peaking at 17.6% in early October 2023 and 12.9% in late December 2023, and was lowest at the end of March 2024. Positivity dropped in February 2024 and remained low until the end of the time period.

For both males and females, COVID-19 episode rates were highest among those aged over 80, followed by those aged 10-19 and 60-69. For those aged over 80, COVID-19 rates were slightly elevated among females compared to males.

Over the study period, the dominant SARS-CoV-2 lineages transitioned from EG.5.1 and its sub-lineages, which were dominant from September to the end of November 2023, to JN.1 and its sub-lineages, which were dominant from December until the end of the study period.

### Severity summary

During this time period, there were 23,838 COVID-19-associated A&E attendances, 45,696 hospital admissions, 2,383 severe hospital admissions and 7,642 COVID-19 deaths across all age groups.

COVID-19-associated A&E attendances, hospital admissions and severe hospital admissions followed a similar trend over the time period, peaking in October 2023 and January 2024, and plateauing from late February 2024 onwards.

The most common COVID-19 outcomes were A&E attendance and hospital admissions. The highest rates for both of these outcomes were seen among infants (under 6 months old) and those aged 80-89 (Table 1). Among infants, the rate of A&E attendances was 713.7 per 100,000 (95%CI: 683.7-744.7) and 1105.4 per 100,000 (95%CI: 1068-1143.8) for hospital admissions. Among those aged 80-89, rates were 231.8 per 100,000 (95%CI: 225.6-238.2) and 536.7 per 100,000 (95%CI: 527.3-546.3), respectively. Absolute numbers of attendances (n=5,348, 22%) and admissions (n=12,383, 27%) remain highest in the 80-89 year olds (Figure 1).

**Table 1.**
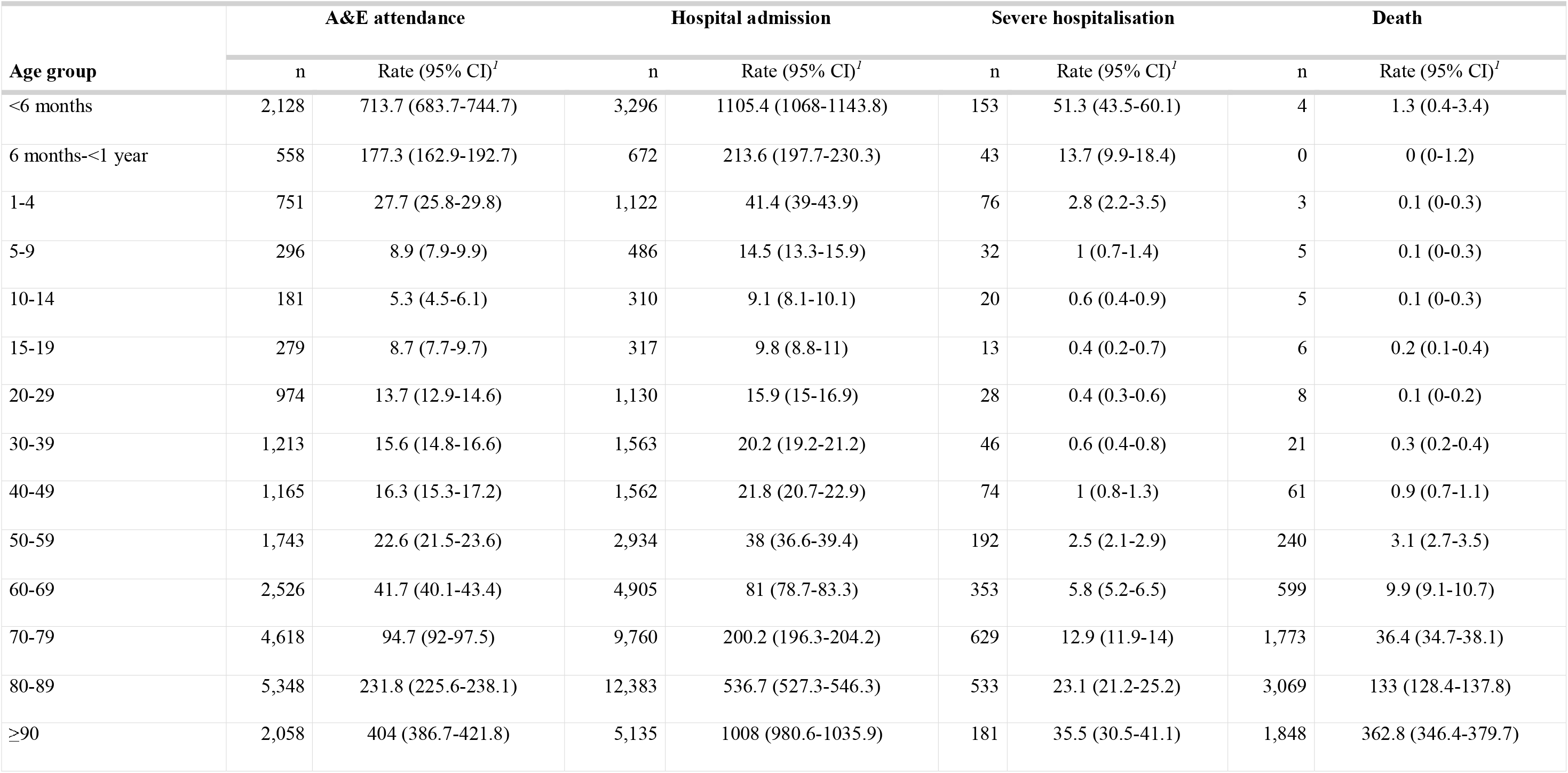
Number and incidence rates per 100,000 of COVID-19 outcomes in England by age group, September 2023 – April 2024.

| Age group | A&E attendance |  | Hospital admission |  | Severe hospitalisation |  | Death |  |
| --- | --- | --- | --- | --- | --- | --- | --- | --- |
|  | n | Rate (95% CI)<sup>I</sup> | n | Rate (95% CI)<sup>I</sup> | n | Rate (95% CI)<sup>I</sup> | n | Rate (95% CI)<sup>I</sup> |
| <6 months | 2,128 | 713.7 (683.7-744.7) | 3,296 | 1105.4 (1068-1143.8) | 153 | 51.3 (43.5-60.1) | 4 | 1.3 (0.4-3.4) |
| 6 months-<1 year | 558 | 177.3 (162.9-192.7) | 672 | 213.6 (197.7-230.3) | 43 | 13.7 (9.9-18.4) | 0 | 0 (0-1.2) |
| 1-4 | 751 | 27.7 (25.8-29.8) | 1,122 | 41.4 (39-43.9) | 76 | 2.8 (2.2-3.5) | 3 | 0.1 (0-0.3) |
| 5-9 | 296 | 8.9 (7.9-9.9) | 486 | 14.5 (13.3-15.9) | 32 | 1 (0.7-1.4) | 5 | 0.1 (0-0.3) |
| 10-14 | 181 | 5.3 (4.5-6.1) | 310 | 9.1 (8.1-10.1) | 20 | 0.6 (0.4-0.9) | 5 | 0.1 (0-0.3) |
| 15-19 | 279 | 8.7 (7.7-9.7) | 317 | 9.8 (8.8-11) | 13 | 0.4 (0.2-0.7) | 6 | 0.2 (0.1-0.4) |
| 20-29 | 974 | 13.7 (12.9-14.6) | 1,130 | 15.9 (15-16.9) | 28 | 0.4 (0.3-0.6) | 8 | 0.1 (0-0.2) |
| 30-39 | 1,213 | 15.6 (14.8-16.6) | 1,563 | 20.2 (19.2-21.2) | 46 | 0.6 (0.4-0.8) | 21 | 0.3 (0.2-0.4) |
| 40-49 | 1,165 | 16.3 (15.3-17.2) | 1,562 | 21.8 (20.7-22.9) | 74 | 1 (0.8-1.3) | 61 | 0.9 (0.7-1.1) |
| 50-59 | 1,743 | 22.6 (21.5-23.6) | 2,934 | 38 (36.6-39.4) | 192 | 2.5 (2.1-2.9) | 240 | 3.1 (2.7-3.5) |
| 60-69 | 2,526 | 41.7 (40.1-43.4) | 4,905 | 81 (78.7-83.3) | 353 | 5.8 (5.2-6.5) | 599 | 9.9 (9.1-10.7) |
| 70-79 | 4,618 | 94.7 (92-97.5) | 9,760 | 200.2 (196.3-204.2) | 629 | 12.9 (11.9-14) | 1,773 | 36.4 (34.7-38.1) |
| 80-89 | 5,348 | 231.8 (225.6-238.1) | 12,383 | 536.7 (527.3-546.3) | 533 | 23.1 (21.2-25.2) | 3,069 | 133 (128.4-137.8) |
| ≥90 | 2,058 | 404 (386.7-421.8) | 5,135 | 1008 (980.6-1035.9) | 181 | 35.5 (30.5-41.1) | 1,848 | 362.8 (346.4-379.7) |

**Figure 1.**
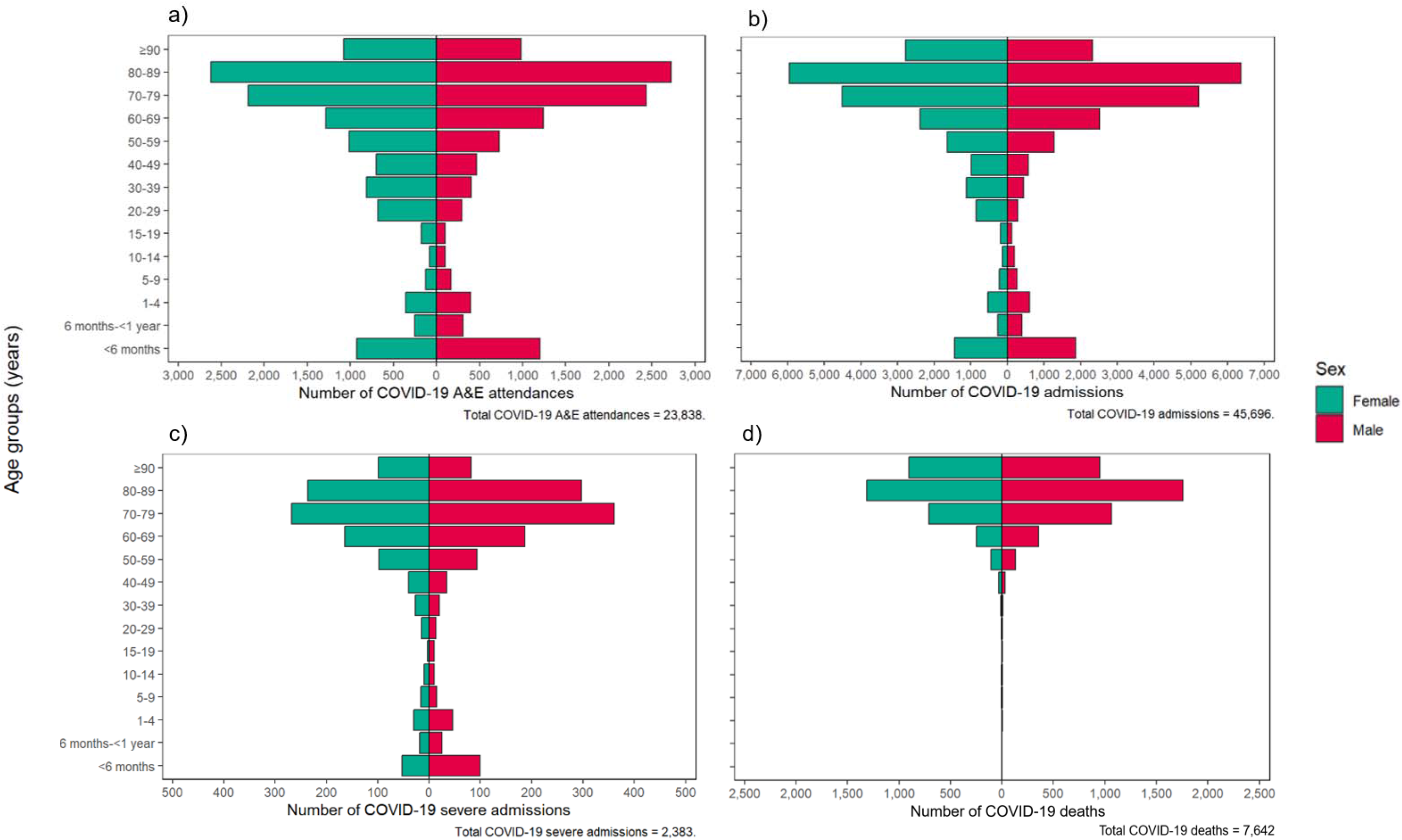
Age-sex pyramid showing number of COVID-19 a) A&E attendances, b) hospital admissions, c) severe hospitalisations, and d) deaths in England, September 2023 – April 2024

The majority of both A&E attendances and hospital admissions among those aged 75 and over were in individuals defined as at clinical risk but not immunosuppressed (70.1% and 69.7% respectively), followed by those who were immunosuppressed (22.8% and 23.7% respectively) (Figure 2). During the time period, 7.1% A&E attendances and 6.6% hospital admissions in those aged 75 and over were among those defined as not at clinical risk.

**Figure 2.**
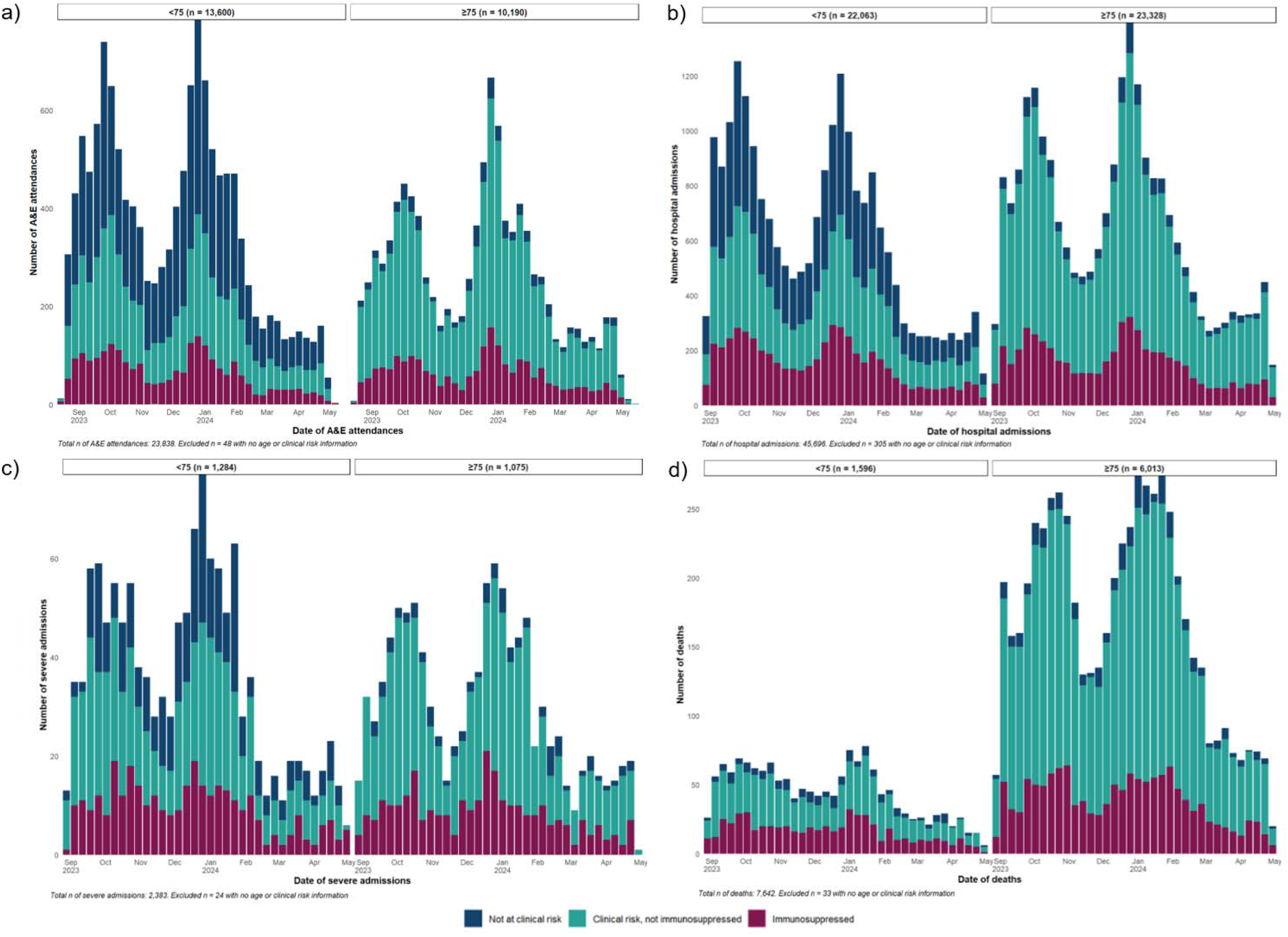
Number of COVID-19 a) A&E attendances, b) hospital admissions, c) severe hospitalisations, and d) deaths by date of event and clinical risk status in England, September 2023 – April 2024

The proportions of COVID-19 A&E attendances and hospital admissions occurring in the immunosuppressed (17.6% and 24.9% respectively) were similar in the under 75 and the 75 years and over age groups. However, much higher proportions of A&E attendances and hospital admissions in the under 75 age group were classified as not at clinical risk (48.9% and 39.3%, respectively) compared to those 75 years and over.

The numbers and rates of severe COVID-19 hospitalisations (length of stay of 2 or more days and admission to intensive care, use of ventilation, or use of oxygen) was lower than A&E attendances and hospital admissions across all age groups. This was also true for rates among infants were also much lower than the other two outcomes with 51.3 per 100,000 (95%CI: 43.5-60.1). Absolute numbers of severe hospitalisations were highest in the 70-79 year olds (n=629).

Similar to A&E attendances and hospital admissions, the majority of severe hospital admissions among those aged 75 and over were of those at clinical risk but not immunosuppressed (65.6%), followed by those who were immunosuppressed (27.4%).

In contrast to the other hospitalisation outcomes, the highest proportion of severe hospitalisations in under 75s were among those who were at clinical risk but not immunosuppressed (46%), followed by those not at clinical risk (28.1%).

Over this time period, 6,013 deaths were observed among those aged 75 and over and 1,596 among those under 75. The rate of COVID-19 deaths was highest among those aged 90 and over (362.8 per 100,000 (95%CI: 346.4-379.7)) followed by those aged 80-89 (133 per 100,000 (95%CI: 128.4-137.8)). For those aged 75 and over, the majority of deaths were among those who were at clinical risk but not immunosuppressed (71.5%), followed by those who were immunosuppressed (22.9%).

In contrast to the hospitalisations outcome, a small proportion of deaths in under 75s were not at clinical risk. The majority of deaths among those aged under 75 were among those who were at clinical risk but not immunosuppressed (52.9%), followed by those immunosuppressed (36.3%).

COVID-19 deaths followed a similar trend to the other outcomes, peaking in October 2023 and January 2024 and plateauing from late February 2024 onwards.

## Discussion

This national-level descriptive study presents a demographic analysis of COVID-19-associated outcomes in England. The number of COVID-19 A&E attendances and hospital admissions were highest among those aged 80-89 year-olds, who are often at higher clinical risk than younger age groups. Most attendances and admissions in those aged under 75 were among those without clinical risk. More severe outcomes, including severe hospitalisation and deaths were primarily seen in those at clinical risk in this age group, who are eligible for vaccination.

Although the rates of most COVID-19 outcomes were highest among those aged 75 and over, high rates of A&E attendance and hospital admission were also observed among infants under 6 months old. This follows evidence that infants under 1 years old are more likely to be more affected by COVID-19, due to lowered immunity and high vulnerability to respiratory illnesses (17). Despite this, lower rates of more severe outcomes such as severe hospitalisations and deaths were observed in this age group. There are currently no vaccines licensed in the England for those aged under 6 months.

Ongoing surveillance of COVID-19 outcomes will continue to inform those most at risk for severe disease, aiding vaccine policy to address those who will benefit most from vaccination.

## Data Availability

Data produced in the present study are available through formal collaboration with
UKHSA. Proposals for collaboration will be considered by the UKHSA Immunisation and Vaccine
Preventable Diseases team to evaluate the scientific quality and feasibility of the proposal.

## Ethics

UKHSA has legal permission, provided by Regulation 3 of The Health Service (Control of Patient Information) Regulations 2002 to process confidential patient information under Section 3(i) (a)–(c), 3(i)(d) (i) and (ii) and 3(iii) as part of its outbreak response activities. This study falls within the research activities approved by the UKHSA Research Ethics and Governance Group.

## Funding

This work was performed as part of UKHSA’s routine work to monitor COVID-19. No external funding received.

## Conflict of interest

The authors declared no conflicts of interest.

## Notes

### Competing Interest Statement

The authors have declared no competing interest.

### Funding Statement

This work was performed as part of UKHSA routine work to monitor COVID-19. No external funding received.

